# Lack of Discernible Benefit of High-Dose Aspirin in the Treatment of Kawasaki Disease

**DOI:** 10.1101/2025.08.03.25332412

**Authors:** Lavina Thadani, Huthaifah Khan, Kwang-Youn Kim, Stanford Shulman, Anne Rowley

## Abstract

In a retrospective single-center cohort study of 460 children with acute-phase Kawasaki Disease, initial treatment with low-dose aspirin with IVIG resulted in similar coronary artery outcomes and IVIG retreatment rates compared to high-dose aspirin with IVIG. These findings support the use of low-dose aspirin for initial management of acute Kawasaki Disease.

## Introduction

Kawasaki disease (KD) is an acute vasculitis of childhood predominantly affecting the coronary arteries (CA).^1^ The goal of acute treatment for KD is to reduce inflammation and arterial damage, and to prevent thrombosis that may occur in damaged coronary arteries. Intravenous immunoglobulin (IVIG) has been demonstrated to improve coronary artery outcomes,^2^ and medium (30-50 mg/kg/day) or high-dose aspirin (80-100 mg/kg/day) continues to be recommended by the American Heart Association in its 2024 update as an adjunctive anti-inflammatory medication.^3^ When the patient’s fever has subsided, a switch to low-dose aspirin (3-5 mg/kg/day) for anti-thrombotic effect is recommended to reduce the possibility of thrombosis in inflamed coronary arteries. However, high-dose aspirin (ASA) can result in adverse effects in children, including gastrointestinal bleeding^4^ and an increased risk of Reye’s syndrome.^5^

Several prior studies in Asian, Israeli, and Canadian populations have questioned the need for medium- or high- dose aspirin for the initial treatment of Kawasaki disease.^6,7,8,9^ These studies showed no significant difference in coronary artery outcomes among patients initially treated with medium- or high-dose aspirin as compared to low-dose aspirin. Some question the need for medium- or high-dose aspirin in acute-phase KD entirely.^10^ An advantage of using low-dose aspirin at diagnosis is the antithrombotic action of aspirin at low doses but not at high doses,^11^ allowing for prevention of thrombosis in aneurysms that may be developing early in the disease process. We found prior studies compelling enough to change our practice for standard treatment for acute KD at our center from IVIG with high-dose ASA to IVIG with low-dose ASA beginning in June 2017. However, the American Heart Association (AHA) guideline on treatment of acute KD remains unchanged in the most recently published guidelines, citing insufficient evidence regarding aspirin dose in acute KD.^3^ In this study, we report outcomes in KD patients treated at our US center at the time of diagnosis with IVIG and high-dose aspirin from 2010 to mid-2017 compared with those treated with IVIG and low-dose aspirin at diagnosis from mid-2017 to 2023. We demonstrate similar coronary artery outcomes and need for IVIG retreatment in the two groups of patients in a previously unstudied US population.

## Methods

### Study Design and Location

We conducted a retrospective cohort study of KD patients treated at Lurie Children’s Hospital between 2010 and 2023. Patients received either high-dose aspirin (80–100 mg/kg/day, 2010–June 2017) or low-dose aspirin (3–5 mg/kg/day, June 2017–2023) with 2 grams/kg of IVIG upon diagnosis. This study was approved by the Lurie Children’s Institutional Review Board.

### Data Collection

Patient data was collected from electronic medical records. General information including age of patient at the time of diagnosis, date of KD diagnosis, patient sex, and baseline blood test values including albumin and C-reactive protein (CRP) were collected to make certain that the patients in the high and low-dose aspirin groups were comparable. We also recorded the date of first IVIG treatment, the number of days between fever onset and first IVIG treatment, the date of second IVIG treatment (if applicable), whether the patient initially received high or low-dose aspirin, and whether or not the patient received corticosteroid therapy. Lastly, using the Boston model,^12^ Z-scores were recorded for the diameter of the right coronary artery (RCA) and left anterior descending coronary artery (LAD) on echocardiography. Baseline Z-scores were obtained from the initial echocardiogram at the time of diagnosis of KD, and maximum Z-scores (Z-max) were obtained from the highest value of all Z-scores for each of the RCA and LAD coronary arteries across all echocardiograms done at diagnosis and in follow-up.

From January 2010 to December 2016, patients with high-risk KD (infants, those with coronary artery abnormalities at baseline, patients presenting in shock) were treated with standard therapy of IVIG and high-dose aspirin. Additional therapies with short course corticosteroids as primary or salvage therapy were administered in a subset of patients in a non-protocolized fashion. From January 2017 – December 2023, adjunctive corticosteroids in a previously reported regimen of tapering corticosteroid therapy^13^ were administered with IVIG and low-dose aspirin to all patients meeting our high-risk criteria.

At our center, the usual reason for retreatment was persistent fever at 36 hours after completion of initial IVIG therapy. In some patients, retreatment was administered because the coronary arteries were enlarging or the CRP level had not decreased after initial therapy. These reasons for retreatment were the same over the entire time period of the study.

### Statistical Analysis

R Statistical Software Version 4.2.0 was used for the statistical analyses. We evaluated the percentage of patients receiving IVIG retreatment, the differences between baseline and maximum Z-scores for the high and low-dose aspirin groups, and the baseline Z-score and Z-max of LAD and RCA across both groups using *t*-tests.

We also performed several subgroup analyses, including assessment of outcomes in patients who did not receive corticosteroid therapy (n=307 of 460), patients with an increase in max Z-score of greater than 1.96 (i.e. 2 standard deviations) from baseline (n=169 of 460), and patients who both had an increase Z-score greater than 1.96 and received corticosteroid therapy (n=95 of 460).

In addition to comparing the change in Z-score from baseline to maximum, we also analyzed outcomes by severity of coronary artery aneurysm (CAA); no CAA=Z-max < 2.5, small CAA= Z-max ≥ 2.5 and < 5, medium CAA= Z-max ≥5 and < 10, and large CAA= Z-max ≥ 10, as defined by the AHA.^14^

## Results

Of the 686 patients diagnosed or treated for KD at Ann & Robert H. Lurie Children’s Hospital of Chicago, 460 patients were included in this study, of which 271 received high-dose aspirin and 189 received low-dose aspirin. For this study, patients were excluded for the following reasons: missing day of diagnosis (n=32), missing day of fever onset (n=5), treated prior to the 4th day of fever onset (n=4) or after the 10th day (n=78), unclear KD diagnosis (n=12), missing baseline or follow-up echocardiogram data (n=80), missing lab data (n=11), or mistake in aspirin dosing (n=4). Of the patients missing echocardiogram data (n=80), 28 patients (15%) were in the low-dose group and 52 patients (19%) were in the high-dose aspirin group.

Patient sex, serum albumin and CRP levels, and days from fever onset to first IVIG dose were similar in the two treatment groups. Patient age was significantly different with the low-dose ASA group being significantly younger than the high-dose ASA cohort (p < 0.001) (Table 1). IVIG retreatment rate in the low-dose aspirin group was 19.6% (37 of 189 patients) compared with 24.0% (65 of 271 patients) in the high-dose group (p = 0.315). In the subgroup of patients who did not receive corticosteroid therapy, the low-dose aspirin group had a 16.0% retreatment rate (15 of 94 patients), while the high-dose group was 19.2% (41 of 213 patients, p = 0.598). Among the patients who developed an increase in Z-max of at least 1.96, or 2 standard deviations from their individual baseline Z-max, the IVIG retreatment rate was 29.4% (20 of 68) in the low-dose aspirin group compared with 31.7% (32 of 101) in the high-dose aspirin group (p = 0.886). In patients who had an increase in Z-max of greater than 2 standard deviations from baseline and who received corticosteroid therapy, 17.4% (4 of 23) of patients in the low-dose aspirin group received IVIG retreatment as compared to 25.0% (18 of 72) patients in the high-dose aspirin group (p=0.639).

**Table 1:** Demographic, clinical, and laboratory data in the two study groups.

|  | Low-Dose | High-Dose | Overall | p-value |
| --- | --- | --- | --- | --- |
| Sex |  |  |  |  |
| Male | 101 (53.4%) | 164 (60.5%) | 265 (57.6%) | 0.157 |
| Female | 88 (46.6%) | 107 (39.5%) | 195 (42.4%) |  |
| Calculated Age at Treatment (months) |  |  |  |  |
| Mean (SD) | 31.6 (26.8) | 42.3 (33.6) | 37.9 (31.4) | <0.001 |
| Infant Status |  |  |  |  |
| Non-Infant | 144 (76.2%) | 226 (83.4%) | 370 (80.4%) | 0.072 |
| Infant | 45 (23.8%) | 45 (16.6%) | 90 (19.6%) |  |
| IVIg Retreatment |  |  |  |  |
| No | 152 (80.4%) | 206 (76.0%) | 358 (77.8%) | 0.315 |
| Yes | 37 (19.6%) | 65 (24.0%) | 102 (22.2%) |  |
| Albumin (g/dL) |  |  |  |  |
| Mean (SD) | 3.40 (0.553) | 3.35 (0.477) | 3.37 (0.510) | 0.241 |
| Initial CRP (mg/L) |  |  |  |  |
| Mean (SD) | 10.3 (8.19) | 11.2 (6.96) | 10.8 (7.49) | 0.251 |
| Days from Fever Onset to First IVIg Treatment |  |  |  |  |
| Mean (SD) | 6.84 (1.72) | 6.93 (1.61) | 6.89 (1.65) | 0.549 |

For most parameters, the two groups were well-matched. Patient age was significantly different between low-dose and high-dose ASA groups.

In addition to comparing IVIG retreatment rates across patients who received low-dose versus high-dose aspirin, we also examined individual patient baseline and maximum Z-scores in each of our subgroups. In the entire population sample, as well as in each of the subgroups, there was no difference between low-dose and high dose-groups in the change from baseline to Z-max for either the RCA or LAD (Table 2).

**Table 2:** Coronary artery outcomes in the two treatment groups, including subgroup analyses.

|  | **All Patients**<br>**n = 460**<br>**189 low-dose**<br>**271 high-dose** | **Subgroup 1**<br>**n = 307**<br>**94 low-dose**<br>**213 high-dose** | **Subgroup 2**<br>**n = 169**<br>**68 low-dose**<br>**101 high-dose** | **Subgroup 3**<br>**n = 95**<br>**23 low-dose**<br>**72 high-dose** |
| --- | --- | --- | --- | --- |
| **RCA Max Z-score** |  |  |  |  |
| Low-dose | 1.52 | 0.89 | 2.12 | 0.72 |
| High-dose | 1.39 | 1.16 | 1.66 | 1.39 |
| **LAD Max Z-score** |  |  |  |  |
| Low-dose | 1.56 | 0.85 | 2.12 | 0.85 |
| High-dose | 1.35 | 1.10 | 1.73 | 1.37 |
| **Overall Max Z-score** |  |  |  |  |
| Low-dose | 2.10 | 1.32 | 2.82 | 1.23 |
| High-dose | 1.89 | 1.60 | 2.33 | 1.96 |
| **RCA Max Z-score minus baseline** |  |  |  |  |
| Low-dose | 0.59 | 0.47 | 1.05 | 0.70 |
| High-dose | 0.69 | 0.66 | 1.22 | 1.22 |
| **LAD Max Z-score minus baseline** |  |  |  |  |
| Low-dose | 0.78 | 0.46 | 2.02 | 1.58 |
| High-dose | 0.65 | 0.56 | 1.64 | 1.52 |
| **Delta Z-max > 1.96 = 1 (%)** |  |  |  |  |
| Low-dose | 36.0% | 24.5% | 100.0% | 100.0% |
| High-dose | 37.3% | 33.8% | 100.0% | 100.0% |

Subgroup 1 - Patients that did not receive treatment with corticosteroids
Subgroup 2 - Patients with an increase between baseline and Zmax of at least 1.96 (2 standard deviations)
Subgroup 3 - Patients that did not receive treatment with corticosteroids and had an increase between baseline and Zmax of at least 1.96 (2 standard deviations)
Max Z-score for LAD and RCA, as well as the average difference in baseline and Z-max for each patient in the low-dose and high-dose ASA groups was similar in the two groups. The data was further split into several subgroups to evaluate possible confounders, yielding no significant differences. All p-values for data shown in this table were greater than 0.05.

In order to further assess CAA outcomes, we examined the overall population and each subgroup based on the degree of their CAA: not present, small, medium, large. Across the overall population and within all subgroups, there was no statistically significant difference between the percentage of children who developed these outcomes in the low-and high-dose aspirin groups.

## Discussion

The most recent 2024 Kawasaki Disease management guideline published by the American Heart Association continues to recommend treatment with medium- or high-dose aspirin at diagnosis of KD, citing a need for more data to support routine use of low-dose aspirin at the time of diagnosis.^3^ We observed similar coronary artery outcomes and need for retreatment in KD patients treated with low-dose aspirin at diagnosis and those treated with higher doses, consistent with prior studies.^6,7,8,9,10^

We did not directly assess length of hospitalization in our study; however, it is unlikely that patients in the low-dose aspirin group experienced prolonged hospital stays, as IVIG retreatment—which generally results in longer hospitalization—was not more frequent in this group. Additionally, while prior studies evaluating aspirin dosing in Kawasaki Disease have been conducted in Canadian ^6^, Middle Eastern ^8,9^, and Asian populations^10^, our study adds new information by evaluating a US-based cohort. Our findings suggest that the lack of benefit of high-dose aspirin is consistent across populations with diverse genetic backgrounds.

Our study is limited by its retrospective nature. Almost all patients that received high-dose aspirin were treated before July 2017. Improvements in echocardiography over time may have more fully delineated CAA, although this should have resulted in more CAA detection in the low-dose aspirin group, which we did not observe. There were more infants in the low-dose group. However, infants are at higher risk of severe outcomes from KD,^15^ and we did not see more severe outcomes in the low-dose group. We began to administer corticosteroid therapy to high-risk KD patients in a tapering regimen in 2017; however, subgroup analyses of KD patients treated with and without corticosteroids did not yield different outcomes in the low- and high-dose aspirin groups. Another unknown confounding factor related to change in care practice over time could have affected results, although we could not identify such a practice change.

## Conclusion

We did not identify any differences in coronary artery outcomes nor IVIG retreatment rates in KD patients treated at diagnosis with low-dose aspirin and IVIG when compared to those treated with high-dose aspirin and IVIG. Use of low-dose aspirin at diagnosis in KD patients should be seriously considered, to reduce potential side effects of high-dose aspirin and to allow for earlier provision of anti-thrombotic therapy in the event that coronary artery complications ensue as a result of KD. This data may inform future updates to KD management guidelines.

## Data Availability

All data produced in the present study are available upon reasonable request to the authors

## Abbreviations

(KD): Kawasaki Disease
(CA): Coronary artery
(IVIG): Intravenous immunoglobulin
(ASA): Aspirin American Heart Association
(RCA): Right coronary artery
(LAD): Left anterior descending
(z-max): Maximum z-score
(CAA): Coronary artery aneurysm

